# Detection of SARS-Cov-2 RNA in serum is associated with increased mortality risk in hospitalized COVID-19 patients

**DOI:** 10.1101/2021.01.14.21249372

**Authors:** Diego A. Rodríguez-Serrano, Emilia Roy-Vallejo, Nelly D. Zurita Cruz, Alexandra Martín Ramírez, Sebastián C. Rodríguez-García, Nuria Arevalillo-Fernández, José María Galván-Román, Leticia Fontán García-Rodrigo, Lorena Vega-Piris, Marta Chicot Llano, David Arribas Méndez, Begoña González de Marcos, Julia Hernando Santos, Ana Sánchez Azofra, Elena Ávalos Pérez-Urria, Pablo Rodriguez-Cortes, Laura Esparcia, Ana Marcos-Jimenez, Santiago Sánchez-Alonso, Irene Llorente, Joan Soriano, Carmen Suárez Fernández, Rosario García-Vicuña, Julio Ancochea, Jesús Sanz, Cecilia Muñoz-Calleja, Rafael de la Cámara, Alfonso Canabal Berlanga, Isidoro González-Álvaro, Laura Cardeñoso, on behalf of the REINMUN-COVID Group

## Abstract

**Background:** COVID-19 has overloaded national health services worldwide. Thus, early identification of patients at risk of poor outcomes is critical. Our objective was to analyse SARS-CoV-2 RNA detection in serum as a severity biomarker in COVID-19.

**Methods and Findings:** Retrospective observational study including 193 patients admitted for COVID-19. Detection of SARS-CoV-2 RNA in serum (CoVemia) was performed with samples collected at 48-72 hours of admission by two techniques from Roche and Thermo Fischer Scientific (TFS). Main outcome variables were mortality and need for ICU admission during hospitalization for COVID-19.

CoVemia was detected in 50-60% of patients depending on technique. The correlation of Ct in serum between both techniques was good (intraclass correlation coefficient: 0.612; p < 0.001). Patients with CoVemia were older (p = 0.006), had poorer baseline oxygenation (PaO2/FiO2; p < 0.001), more severe lymphopenia (p < 0.001) and higher LDH (p < 0.001), IL-6 (p = 0.021), C-reactive protein (CRP; p = 0.022) and procalcitonin (p = 0.002) serum levels.

We defined “relevant CoVemia” when detection Ct was < 34 with Roche and < 31 for TFS. These thresholds had 95% sensitivity and 35 % specificity. Relevant CoVemia predicted death during hospitalization (OR 9.2 [3.8 − 22.6] for Roche, OR 10.3 [3.6 − 29.3] for TFS; p < 0.001). Cox regression models, adjusted by age, sex and Charlson index, identified increased LDH serum levels and relevant CoVemia (HR = 9.87 [4.13-23.57] for TFS viremia and HR = 7.09 [3.3-14.82] for Roche viremia) as the best markers to predict mortality.

**Conclusions:** CoVemia assessment at admission is the most useful biomarker for predicting mortality in COVID-19 patients. CoVemia is highly reproducible with two different techniques (TFS and Roche), has a good consistency with other severity biomarkers for COVID-19 and better predictive accuracy.

**AUTHOR SUMMARY:** COVID-19 shows a very heterogeneous clinical picture. In addition, it has overloaded national health services worldwide. Therefore, early identification of patients with poor prognosis is critical to improve the use of limited health resources. In this work, we evaluated whether baseline SARS-CoV2 RNA detection in blood (CoVemia) is associated with worse outcomes. We studied almost 200 patients admitted to our hospital and about 50-60% of them showed positive CoVemia. Patients with positive CoVemia were older and had more severe disease; CoVemia was also more frequent in patients requiring admission to the ICU. Moreover, we defined “relevant CoVemia”, as the amount of viral load that better predicted mortality obtaining 95% sensitivity and 35% specificity. In addition, relevant CoVemia was a better predictor than other biomarkers such as LDH, lymphocyte count, interleukin-6, and indexes used in ICU such as qSOFA and CURB65.

In summary, detection of CoVemia is the best biomarker to predict death in COVID-19 patients. Furthermore, it is easy to be implemented and is reproducible with two techniques (Roche and Thermo Fisher Scientific) that are currently used for diagnosis in nasopharyngeal swabs samples.

## INTRODUCTION

The wide spectrum of COVID-19 severity ranges from asymptomatic to critical cases, albeit less than 10% of patients develop a severe disease [1]. Even though only a minority of patients need hospitalization for COVID-19, the higher transmission rate of SARS-Cov-2 compared to other viruses, the absence of previous immunity in the population, and the high incidence of this disease in a short period of time are collapsing health care systems worldwide [2]. One remaining challenge from COVID-19 is the difficulty of predicting individual prognosis since determinants of disease severity remain unclear. Previous studies have suggested that age, male sex, obesity, hypertension, and underlying diseases like hematologic malignancies are associated with worse prognosis [3–5]. Likewise, some blood biomarkers are able to predict the emergence of the cytokine storm leading to severe acute respiratory syndrome, the most frequent cause of clinical deterioration in COVID-19 patients [6–8].

Despite the tropism of SARS-CoV-2 for the upper respiratory tissue [9], the relevance of its viral load in nasopharyngeal samples remains controversial [10]. However, several authors have reported the detection of SARS-CoV-2 RNA in serum or plasma samples (henceforth CoVemia) associated with a worse prognosis, assessed as higher probability of clinical deterioration, higher levels of interleukin (IL)-6, IL-5 or CXCL10, intensive care unit (ICU) admission, critical disease and death [8,11–13]. Notwithstanding, the detection of CoVemia with more sensitive techniques was not associated with mortality, but with immune suppression status [14].

Therefore, we aimed to assess whether there is an association of CoVemia with COVID-19 severity using two different real-time reverse-transcription polymerase chain reaction (rRT-PCR) techniques,and compare them with other suggested severity biomarkers.

## RESULTS

### Demographic and clinical characteristics of the study population

One hundred and ninety-three patients were included; their main demographic and baseline clinical characteristics and laboratory findings are shown in Table 1 and S1 Table.

**Table 1.** Baseline clinical characteristics of patients according to SARS-CoV-2 RNA detection in blood by Roche technique

|  | Study Population<br>(n = 193) | Absent<br>(n = 100) | CoVemia<br>Present<br>(n = 93) | P<br>Value |
| --- | --- | --- | --- | --- |
| **Age** | **63 (55 - 71)** | **61 (54 - 68)** | **67 (59 - 73)** | **0.001** |
| Male sex | 134 (69) | 68 (68) | 66 (71) | 0.655 |
| Comorbidities | 137 (71) | 67 (67) | 70 (75) | 0.206 |
| Duration of symptoms<br>at admission (days) | 6 (4 - 8) | 7 (5 - 9) | 6 (4-8) | 0.120 |
| **Baseline PaO<sub>2</sub>/FiO<sub>2</sub>** | **188 (100 - 282)** | **250 (145 - 348)** | **146 (90 - 214)** | **< 0.001** |
| *Treatment during*<br>*hospitalization* |  |  |  |  |
| Hydroxychloroquine | 186 (96) | 97(97) | 89(96) | 0.629 |
| Lopinavir/Ritonavir | 168 (87) | 83 (83) | 85 (91) | 0.083 |
| Azithromycin | 133 (69) | 69 (69) | 64 (69) | 0.978 |
| Interferon-β | 8 (4) | 2 (2) | 6 (6) | 0.121 |
| **Glucocorticoids** | **134 (69)** | **62 (62)** | **72 (77)** | **0.020** |
| **Methylprednisolone**<br>**bolus** | **101 (63)** | **42 (53)** | **59 (72)** | **0.014** |
| **Tocilizumab** | **91 (47)** | **36 (36)** | **55 (59)** | **0.001** |
| *Laboratory Findings* |  |  |  |  |
| WBC (10<sup>3</sup>/mm<sup>3</sup>)<br>NR: 4.00 - 10.00 | 7.18 (4.82 - 9.61) | 6.89 (4.66 - 9.35) | 7.47 (5.00 - 10.36) | 0.374 |
| **Lymphocytes/mm<sup>3</sup>** | **790 (590 - 1165)** | **950 (720 - 1270)** | **660 (510 - 860)** | **< 0.001** |
| NR: 1.00 -4.00 |  |  |  |  |
| Creatinine (mg/dl) | 0.86 (0.67 – 1.09) | 0.85 (0.69 – 1.03) | 0.90 (0.63 – 1.16) | 0.563 |
| NR:0.70 - 1.20 |  |  |  |  |
| **LDH (U/L)** | **390 (278 - 512)** | **314 (246 - 428)** | **461 (371 - 565)** | **< 0.001** |
| NR: 135 - 225 |  |  |  |  |
| CK (U/L) | 100 (49 - 270) | 94 (49 - 155) | 110 (56 - 336) | 0.315 |
| NR: 20 - 180 |  |  |  |  |
| **Serum IL-6 (pg/ml)** | **20.7 (7.9 – 52.1)** | **16.4 (7.5 – 41.3)** | **27.0 (7.9 – 70.3)** | **0.031** |
| NR: < 30 pg/ml |  |  |  |  |
| Ferritin (ng/ml) | 1542 (871 – 2617) | 1418 (717 - 2068) | 1637 (944 - 3088) | 0.267 |
| NR: 30 - 400 |  |  |  |  |
| **CRP (mg/dL)** | **12.2 (5.5 - 23.0)** | **10.3 (5.1 – 19.2)** | **14.7 (7.5 – 25.7)** | **0.026** |
| NR: 0.00 - 0.50 |  |  |  |  |
| **PCT (ng/ml)** | **0.19 (0.11 – 0.41)** | **0.13 (0.08 – 0.31)** | **0.26 (0.14 – 0.56)** | **0.002** |
| NR: 0.05 - 0.09 |  |  |  |  |
| **D-dimer (µg/ml)** | **0.78 (0.52 – 1.47)** | **0.7 (0.45 – 1.22)** | **0.87 (0.67 – 1.75)** | **0.030** |
| NR:0.14 - 0.50 |  |  |  |  |

All categorical variables are expressed as number (%) and quantitative variables as median (p25-75).
PaO<sub>2</sub>/FiO<sub>2</sub>: arterial oxygen tension – fraction of inspired oxygen ratio; WBC: White Blood Count; NR:
Normal Range; LDH: Lactate Dehydrogenase; CK: Creatin Kinase; IL6: Interleukin-6; CRP: C-Reactive
Protein; PCT: Procalcitonin.

**S1 Table.**
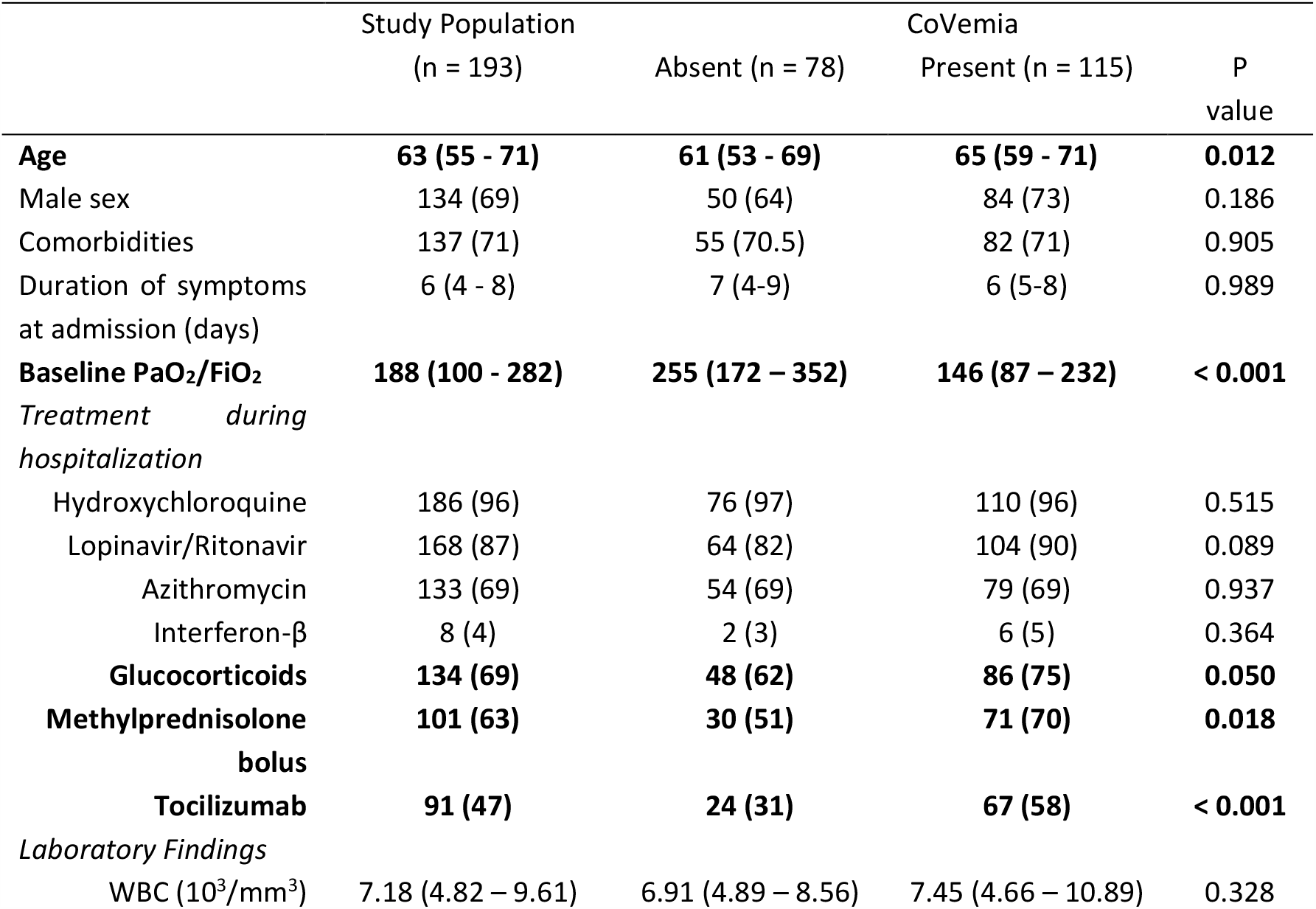
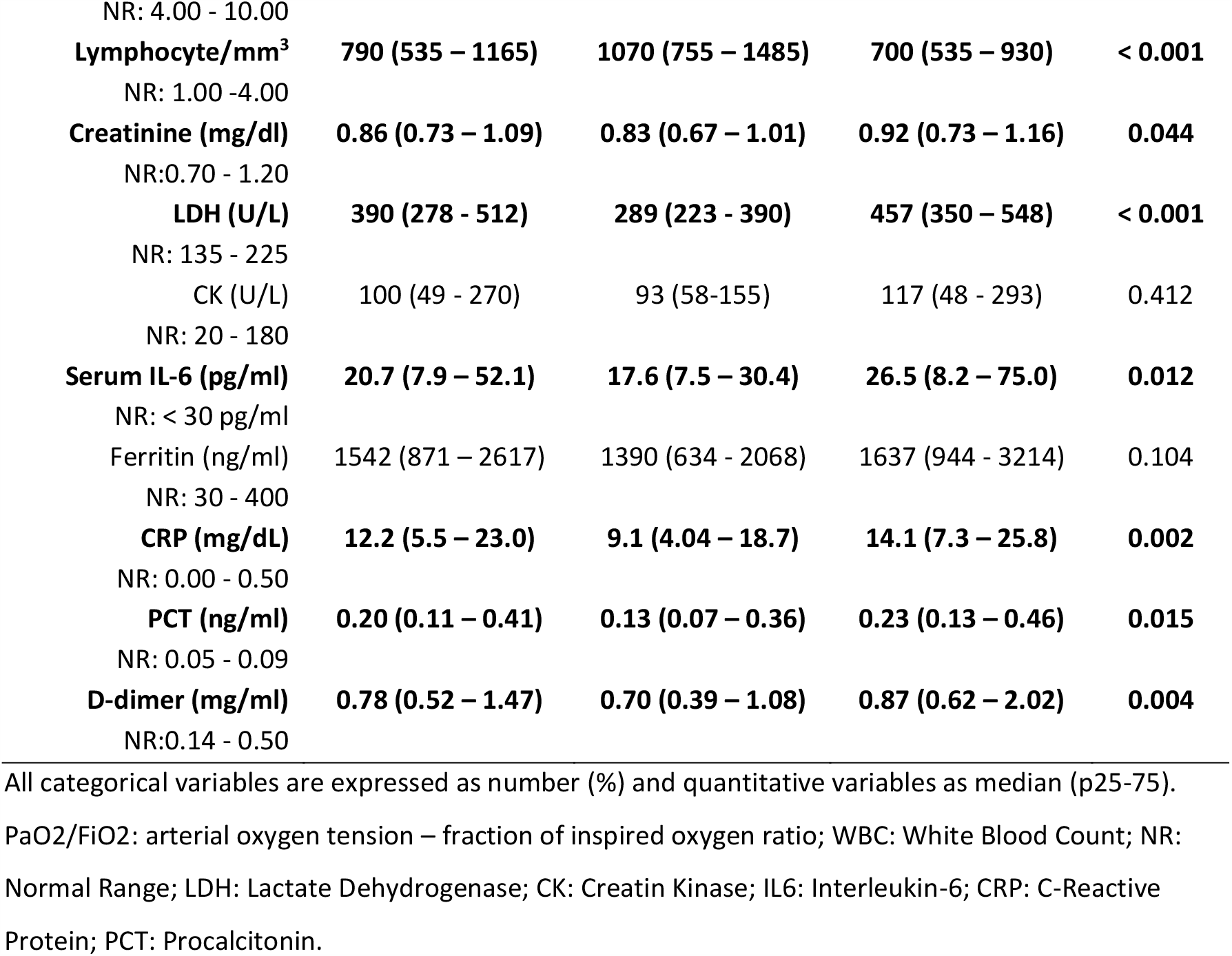
Baseline clinical characteristics of patients according to SARS-CoV-2 RNA detection in blood by Thermo Fisher Systems technique.

Depending on the technique, CoVemia was detected in 95 (48%; Roche) and 117 (59%; TFS) patients; the correlation between Ct (Cycle threshold) in serum obtained with both techniques was very good (r = 0.738 [0.667 − 0.795], p < 0.001), and detection by TFS was more sensitive than by Roche (S1A Figure). The agreement to detect CoVemia was 75.5% and the intraclass correlation coefficient was 0.612 (p < 0.001). Conversely, the correlation of Ct between nasopharyngeal and throat swab (NPTS) and serum samples was weak either with TFS or Roche techniques (S1B and 1C Figure, respectively).

**S1 Figure:**
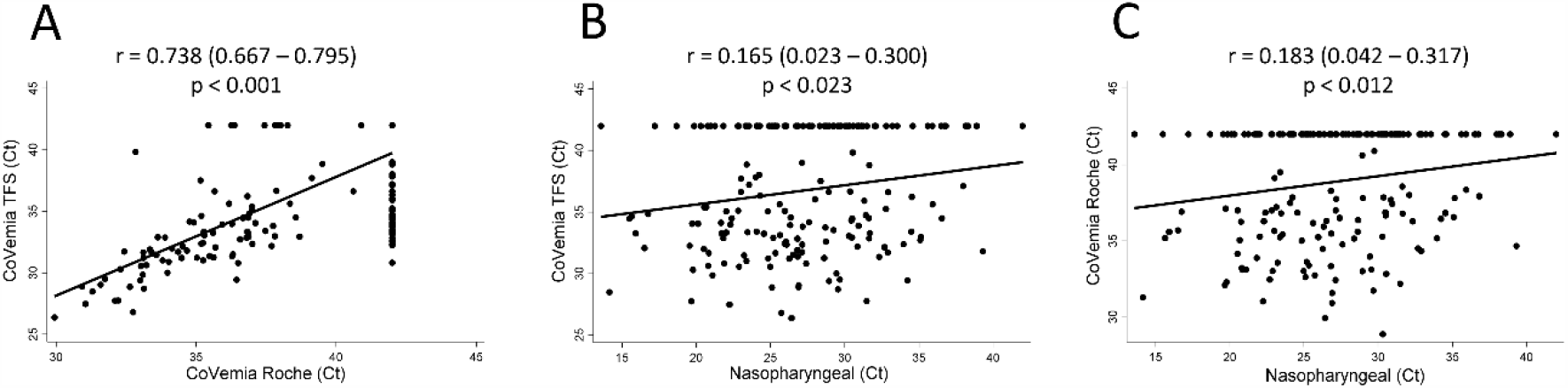
Correlation between quantitative detection of SARS-CoV-2 in serum (CoVemia), either with Thermo Fisher Scientific (TFS) or Roche, and in nasopharyngeal and throat swab (NPTS). A) Correlation between CoVemia with TFS and Roche techniques; Correlation between SARS-CoV-2 detection in NPTS samples and Covemia using the TFS technique (B) or the Roche technique (C). Data are shown as dot plots of the mean Ct values of the RT-PCR for SARS-CoV-2 and the fitted linear prediction obtained with the option *lfit* of the command *twoway* of Stata. Correlation coefficients and significance levels were estimated with the Pearson’s test.

Patients with detectable CoVemia were older (p = 0.006), with worse baseline arterial oxygen tension/fraction of inspired oxygen ratio (PaO_2_/FiO_2_; p < 0.001), lower lymphocyte count (p < 0.001) and higher LDH (p < 0.001), IL-6 (p = 0.021), C-reactive protein (CRP; p = 0.022) and procalcitonin (p = 0.002) serum levels compared to patients without detectable CoVemia. In addition, they were more frequently treated with glucocorticoids (p = 0.016), either oral or bolus (p = 0.015), and with tocilizumab (p = 0.001) (Table 1 and S1 Table).

There were no significant differences in symptom duration at the time of sample collection between patients with and without CoVemia (9 days [95%CI: 4 – 13] and 8 [6 – 11] respectively; p = 0.223) with both techniques. Neither we observed correlation between disease duration and viral RNA detection in serum with both techniques (r = − 0.048 [-0.188 − 0.094], p = 0.510 for Roche, and r = −0.104 [-0.241 − 0.038], p = 0.151 for TFS).

### Prevalence of SARS-CoV-2 viremia is higher in patients requiring ICU admission

In order to evaluate the relationship between COVID-19 severity and viremia we first studied the relative frequency of CoVemia in patients who required ICU admission and those who did not. As shown in Figure 1, viremia was significantly more frequent for both techniques in patients requiring ICU (p < 0.001 for Roche and p = 0.002 for TFS). Although TFS was more sensitive to detect CoVemia, it seemed to be less specific to predict the need for ICU (OR 2.47 [95% CI: 1.37 − 4.47], p = 0.003 for TFS vs 3.04 [1.71 − 5.44] p < 0.001 for Roche).

**Fig. 1.**
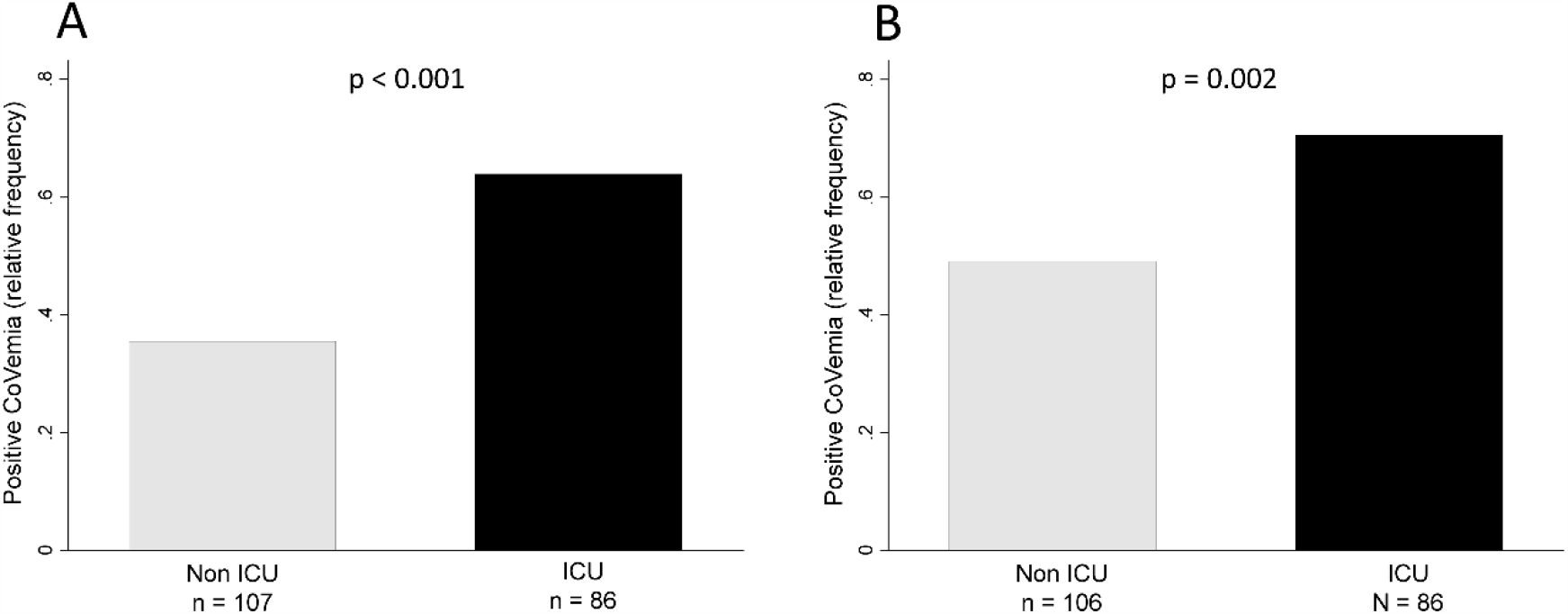
Detection of SARS-CoV-2 RNA in serum (CoVemia) is more frequent in patients requiring intensive care unit (ICU) admission. A) Roche technique. B) Thermo Fisher Scientific technique. Statistical significance was determined with the Chi squared test.

Additionally, we performed a multivariable logistic regression in order to determine whether CoVemia can help predicting ICU admission compared to other variables. The best model to predict the need for ICU admission included the following variables: COPD, total lymphocyte count and PaO_2_/FiO_2_ at admission (Table 2). When we forced CoVemia into this model, it did not reach statistical significance either when it was assessed by Roche or by TFS (Table 2).

**Table 2.**
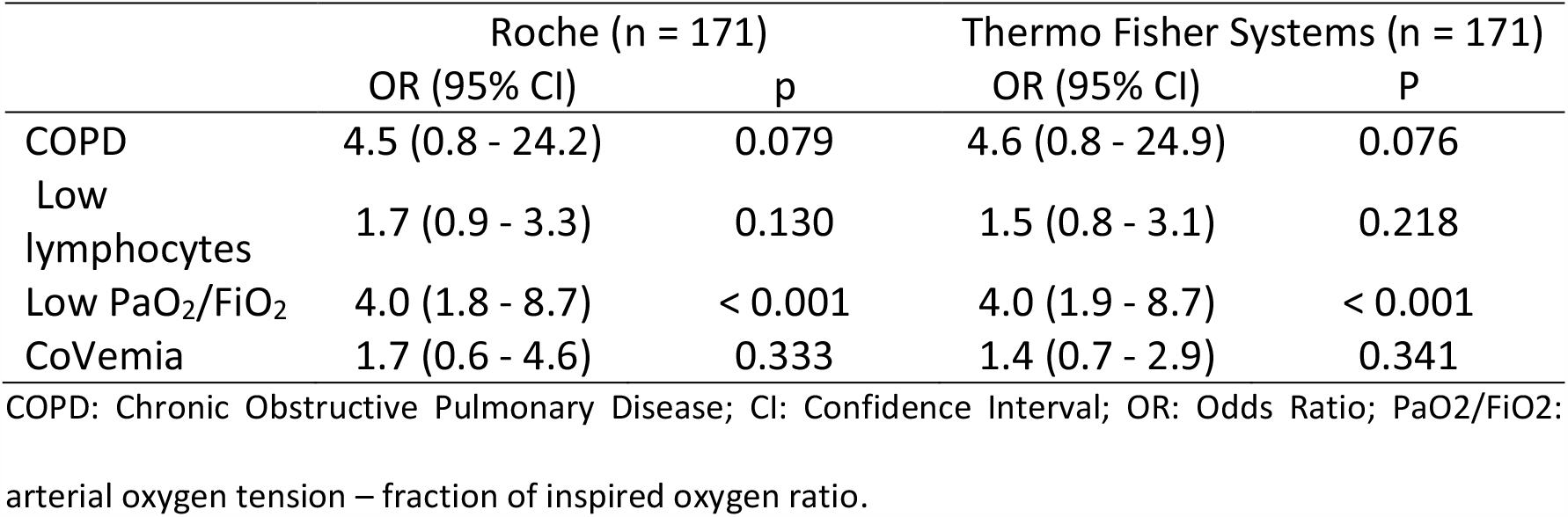
Variables predicting the need for Intensive Care Unit admission

In addition, when we specifically studied the 86 patients that required ICU admission, no association was found between the presence of CoVemia and the development of multiorgan dysfunction syndrome prior to admission and during the first 24 hours at the ICU (S2 Table).

**S2 Table.** Multiorgan failure and dysfunction syndrome according to baseline SARS-CoV-2 RNA detection in serum (Thermo Fisher Scientific).

|  | Negative CoVemia<br>(n = 26) | Positive CoVemia<br>(n = 60) | Total<br>(n = 86) | P value |
| --- | --- | --- | --- | --- |
| **MODS (pa)** | 12 (46.2) | 26 (43.3) | 38 (44.2) | 0.983 |
| **MODS (24 hrs)** | 19 (73.1) | 40 (66.7) | 59 (68.6) | 0.832 |
| **MOF (pa)** | 1 (3.8) | 3 (5.0) | 4 (4.7) | > 0.99 |
| **MOF (24 hrs)** | 13 (50) | 29 (48.3) | 42 (48.8) | 0.922 |
| **Cardiovascular (pa)** | 1 (3.8) | 5 (8.3) | 6 (7.0) | 0.661 |
| **Liver (pa)** | 3 (11.5) | 4 (6.7) | 7 (8.1) | 0.674 |
| **Renal (pa)** | 5 (19.2) | 9 (15.0) | 14 (16.3) | 0.758 |
| **CNS (pa)** | 2 (7.7) | 6 (10.0) | 8 (9.3) | > 0.99 |
| **Coagulation (pa)** | 4 (15.4) | 12 (20.0) | 16 (18.6) | 0.767 |
| **Cardiovascular (24 hrs)** | 14 (50.0) | 33 (55.0) | 47 (54.7) | 0.719 |
| **Liver (24 hrs)** | 2 (7.7) | 6 (10.0) | 8 (9.3) | > 0.99 |
| **Renal (24 hrs)** | 4 (15.4) | 9 (15.0) | 13 (15.1) | > 0.99 |
| **CNS (24 hrs)** | 2 (7.7) | 3 (5.0) | 5 (5.8) | 0.648 |
| **Coagulation (24 hrs)** | 6 (23.1) | 9 (15.0) | 15 (17.4) | 0.435 |

MODS (pa): Multiorgan Dysfunction syndrome prior to admission; MODS (24 hrs): Multiorgan Dysfunction syndrome during the first 24 hours at the ICU; MOF (pa): Multiorgan Failure prior to admission; MOF (24 hrs): Multiorgan Failure during the first 24 hours at the ICU. Cardiovascular (pa): cardiovascular
dysfunction/failure prior to admission. Liver (pa): liver dysfunction/failure prior to admission; Renal (pa): renal dysfunction/failure prior to admission; CNS (pa): central nervous system dysfunction/failure prior to admission; Coagulation (pa): platelets dysfunction/failure prior to admission; Cardiovascular (24 hrs): cardiovascular dysfunction/failure during the first 24 hours at the ICU; Liver (24 hrs): dysfunction/failure during the first 24 hours at the ICU; Renal (24 hrs): dysfunction/failure during the first 24 hours at the ICU; CNS (24 hrs): central nervous system dysfunction/failure during the first 24 hours at the ICU; Coagulation (24 hrs): platelets dysfunction/failure during the first 24 hours at the ICU.

### SARS-CoV-2 viremia predicts mortality in COVID-19 admitted patients

As it was previously suggested that CoVemia can be associated with a higher risk of mortality, we also analyzed how both techniques predicted death during admission. TFS was more sensitive than Roche’s technique but less specific either in the whole population (OR 5.8 [2.6 − 13.3] for Roche and 3.3 [1.4 − 7.4] for TFS), or after stratification by requirement of ICU (S2 Figure).

**S2 Figure.**
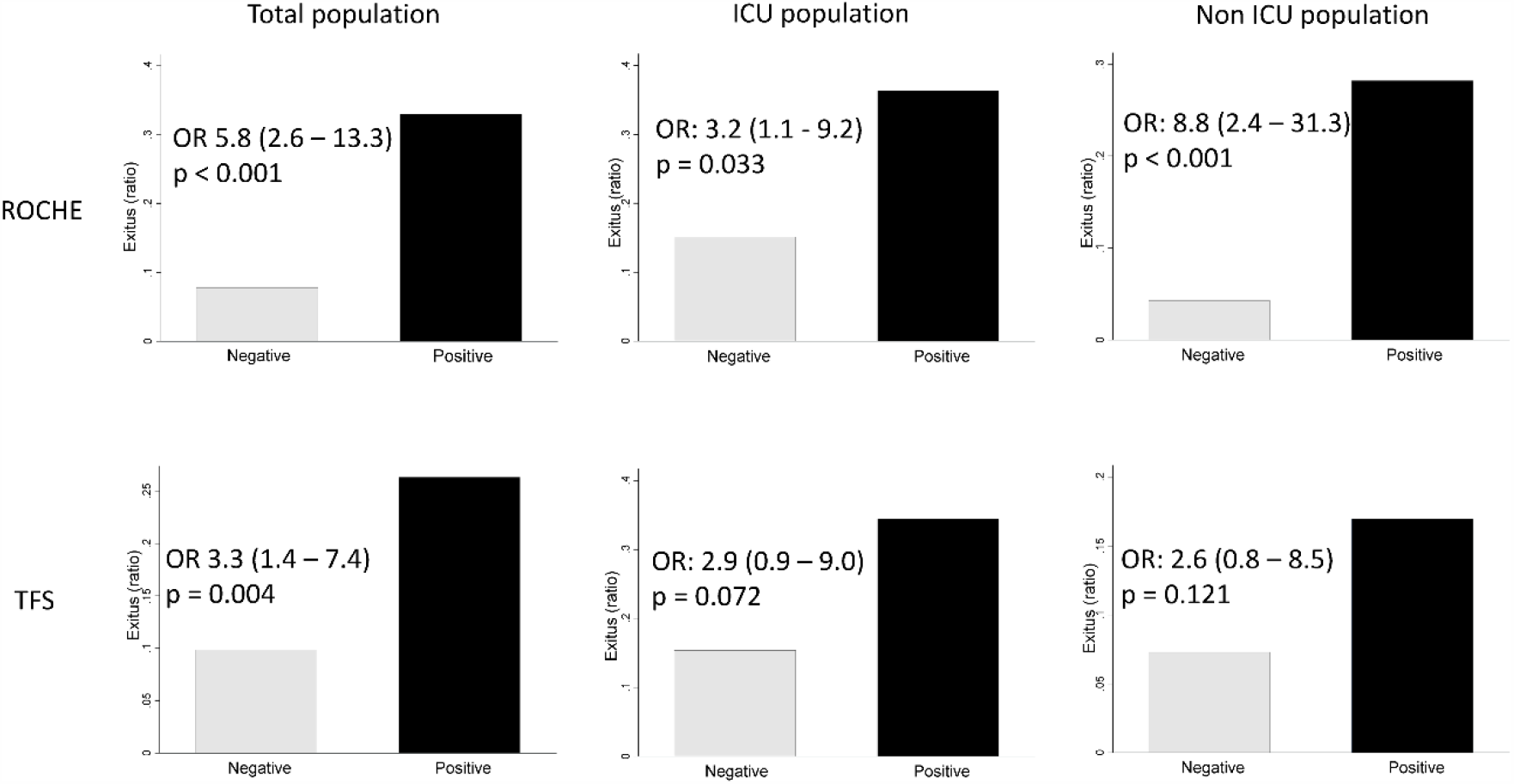
Mortality is higher among patients with positive CoVemia, either in the whole population (left panels), in those patients requiring ICU admission (middle panels) or those no requiring ICU (right panels). Odds ratio (OR) and significance level for mortality according to the presence of CoVemia was estimated with the *cs* command of Stata. TFS: Thermo Fisher Scientific.

Therefore, we analyzed whether a threshold of CoVemia (as Ct) could help to better predict mortality during hospitalization. As shown in Figure 2A and 2B the area under ROC curves for Roche and TFS were quite similar (0.736 [0.642 − 0.829] vs 0.702 [0.603 − 0.801] respectively; p = 0.323). The best cut-off to predict mortality was 34 Ct for Roche (sensitivity 91%, specificity 38%) and 31 Ct for TFS (sensitivity 93%, specificity 32%). Using these cut-off points to define “relevant CoVemia”, the capability to predict mortality improved with both techniques: OR 9.2 (3.8 − 22.6) for Roche and 10.3 (3.6 − 29.3) for TFS (Figures 2C and 2D, respectively).

**Fig. 2.**
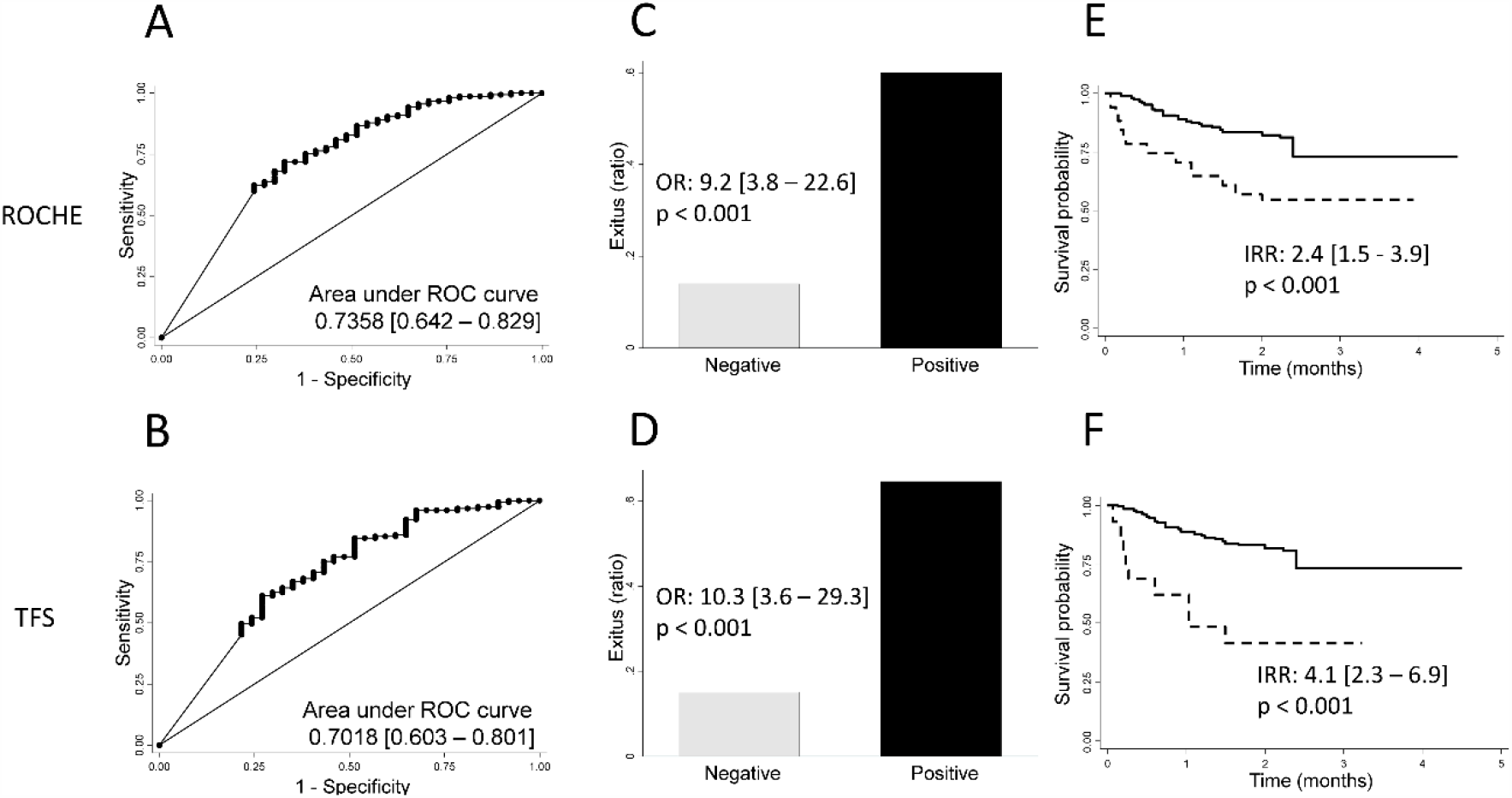
Comprehensive analysis of CoVemia as a prognostic marker of mortality in patients hospitalized for severe COVID-19. ROC curve-analysis for mortality prediction with Ct values in serum of all patients, according to Roche (A) and Thermo Fisher Scientific [TFS] (B) techniques. Proportion of deceased patients according to relevant CoVemia determined by Roche (C) and TFS (D) techniques. Survival analysis with Kaplan-Meier estimator of patients hospitalized for COVID-19 who presented (dotted lines) and patients who did not present (solid lines) relevant CoVemia according to Roche (E) and TFS (F) techniques.

In addition, Kaplan-Meier curves confirmed that those patients with “relevant CoVemia” at admission survived less than those with non-relevant detection of SARS-CoV-2 RNA (Figures 2E and 2F), being slightly higher the incidence risk ratio for CoVemia determined by TFS compared to Roche (IRR 4.1 [2.3 − 6.9] vs 2.4 [1.5 − 3.9] respectively).

### SARS-CoV-2 viremia correlates with severity biomarkers previously described in COVID-19 patients

In order to further validate the usefulness of CoVemia as predictor of mortality in COVID-19, we analyzed its association with clinical and laboratory parameters that have been previously associated with worse outcomes [2,7,15]. As shown in Figure 3, higher levels of CoVemia were more frequent in elderly patients (p = 0.011) and correlated with higher qSOFA (p = 0.022), CURB65 (p = 0.014) and Charlson comorbidity (p = 0.078) indexes.

**Fig. 3.**
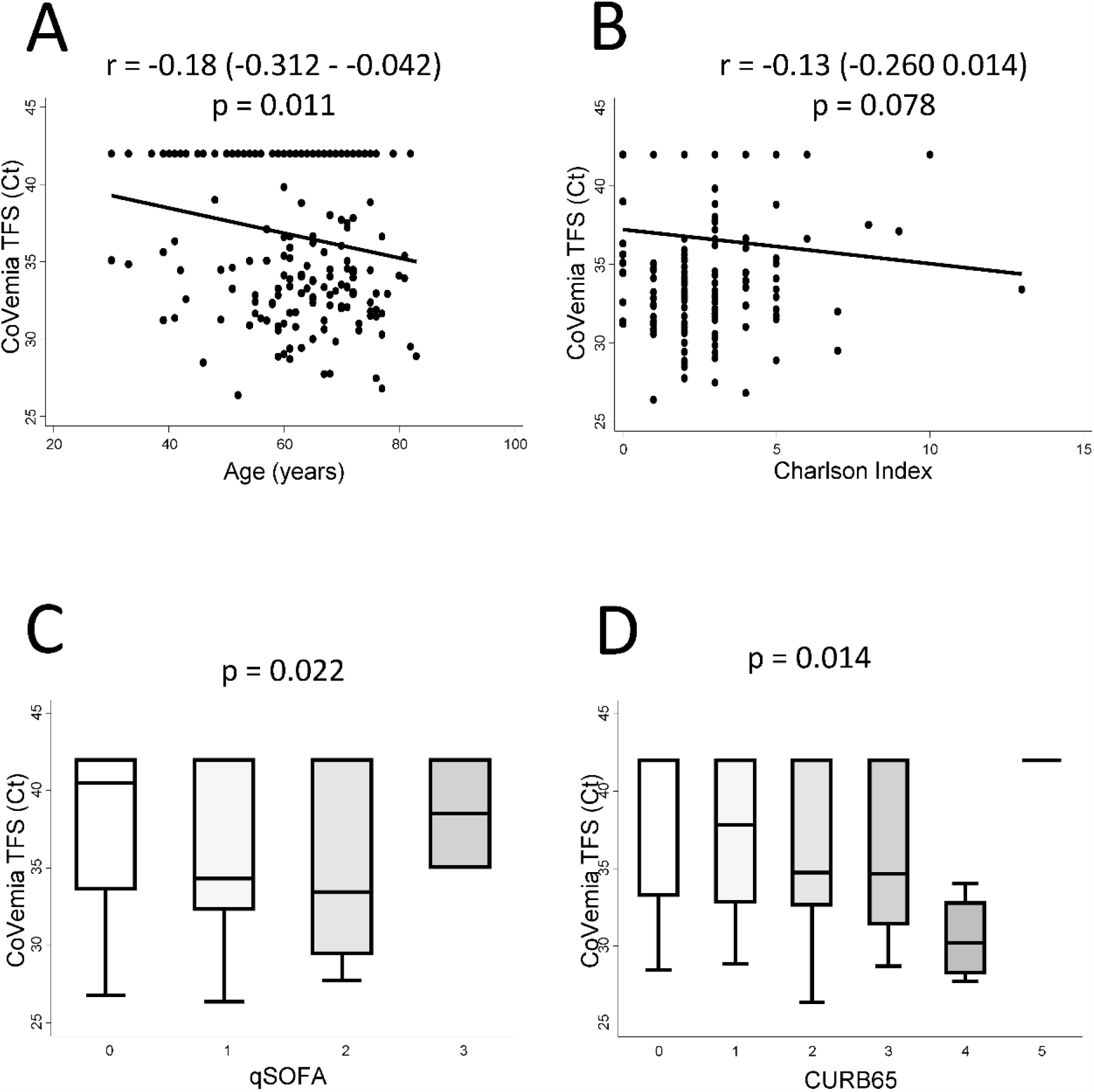
Association between CoVemia and main sociodemographic and clinical characteristics of patients hospitalized with severe COVID-19. Correlation between CoVemia levels (Ct value) and (A) age (years) and (B) Charlson comorbidity index (score). Distribution of CoVemia levels according to qSOFA index (C), and CURB65 scale (D) scores. All Ct data shown here were obtained by rRT-PCR in serum with Thermo Fisher Scientific (TFS) technique. In A and B panels data are shown as dot plots of the corresponding values and the fitted linear prediction (black line) obtained with the option *lfit* of the command *twoway* of Stata. Correlation coefficients and significance levels were estimated with the Pearson’s test. In C and D panels data are shown as box-plot representing the interquartile range (p75 upper edge, p25 lower edge, p50 midline in the box), p95 (line above the box), and p5 (line below the box). Significance level was determined with the Kruskall Wallis test.

Regarding laboratory parameters, CoVemia showed correlation with IL-6 serum levels and lymphocyte count (Figures 4A and 4B; p = 0.014 and p < 0.001, respectively), and a better correlation with lactate dehydrogenase serum levels (Figure 4C; p < 0.001). However, lower significant association was observed with ferritin and D-dimer levels (Figures 4D and 4E; p = 0.073 and p = 0.023, respectively).

**Fig. 4.**
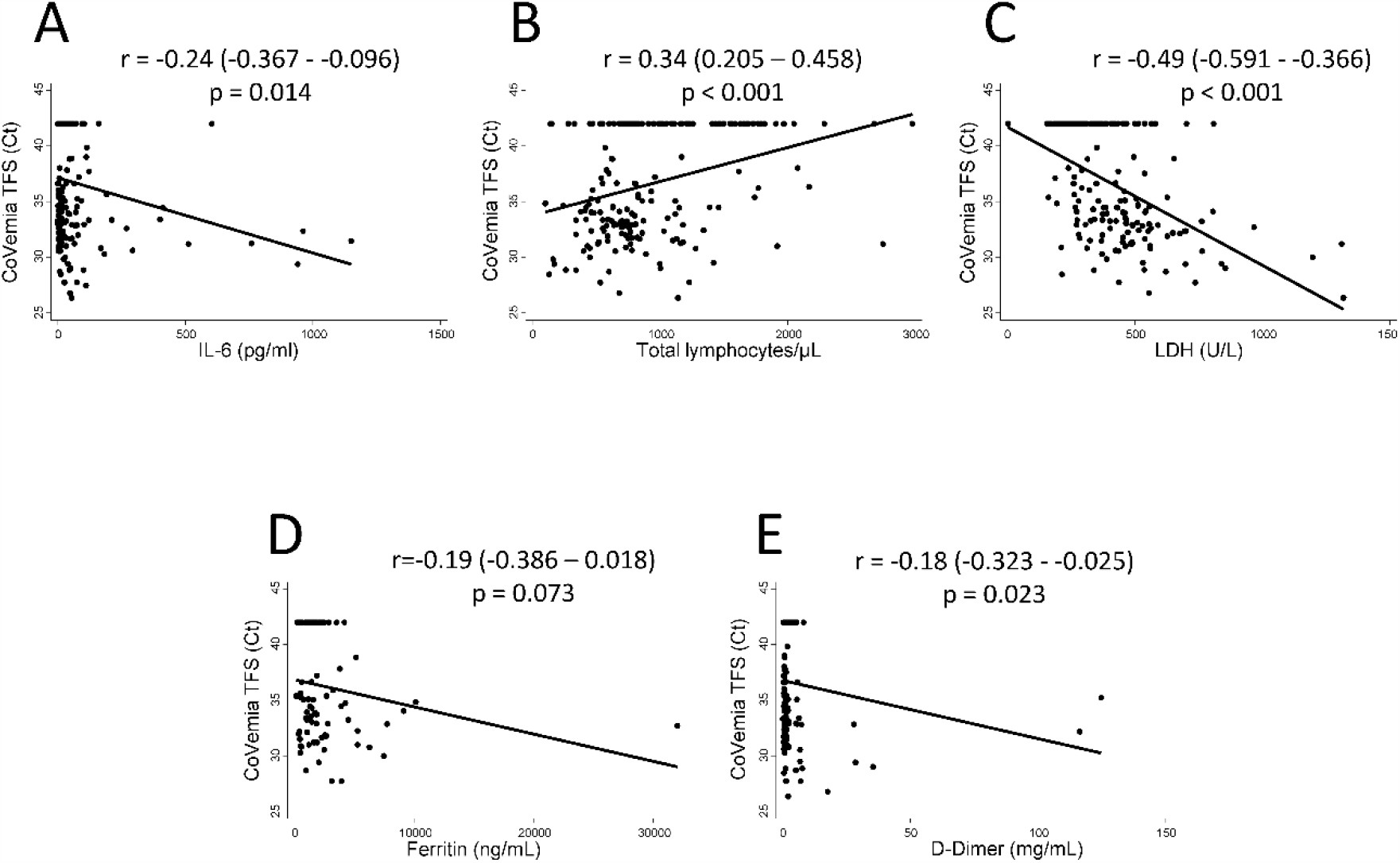
CoVemia correlates with the main laboratory prognosis biomarkers described in patients hospitalized with severe COVID-19. Correlation between CoVemia levels (Ct value by Thermo Fisher Scientific [TFS] technique) and IL-6 levels (pg/mL) (A), lymphocytes count (cel/mm3) (B), LDH levels (U/L) (C), ferritin levels (ng/mL) (D), D-Dimer levels (mg/mL) (E). Data are shown as dot plots of the corresponding values and the fitted linear prediction (black line) obtained with the option *lfit* of the command *twoway* of Stata. Correlation coefficients and significance levels were estimated with the Pearson’s test.

Finally, we analyzed the hazard ratio for mortality of CoVemia in comparison with all these parameters. All models were adjusted by age and Charlson index, which significantly affected survival of COVID-19 patients, as well as by sex, which did not significantly affect survival. Baseline qSOFA and CURB65 showed the worst capability to predict survival (Table 3, model 1). On the contrary, the best predictors always included baseline *“relevant CoVemia”* and high LDH serum levels (Table 3, all models). Interestingly, even when relevant CoVemia, and high LDH levels were included together in model 4, both were able to independently predict survival, although the hazard ratio was higher for the former (Table 3 last two columns; HR 9.87 [4.13 − 23.57] for TFS CoVremia and 7.09 [3.39 − 14.82] for Roche CoVemia; HRs for high LDH 2.48 [1.21 − 5.09] with TFS CoVemia, and 2.39 [1.17 − 4.90] with Roche CoVemia).

**Table 3.** Mortality predictive value of CoVemia compared to other factors

|  | Model 1 |  | Model 2 |  | Model 3 |  | Model 4 |  |
| --- | --- | --- | --- | --- | --- | --- | --- | --- |
|  | HR | p | HR | p | HR | p | HR | P |
| qSOFA | 0.92 | N.S. | N.I. | - | N.I. | - | N.I. | - |
| CURB65 | 1.06 | N.S. | N.I. | - | N.I. | - | N.I. | - |
| Low Ly. | 1.85 | 0.080 | - | - | 1.85 | 0.091 | 1.37 | 0.388 |
| High IL-6 | 1.95 | 0.055 | 1.74 | 0.121 | - | - | 1.45 | 0.304 |
| High LDH | 2.74 | 0.005 | 2.74 | 0.008 | 2.57 | 0.011 | - | - |
| CoVemia TFS | 13.5 | < 0.001 | 11.30 | < 0.001 | 10.4 | < 0.001 | 9.87 | < 0.001 |
| CoVemia Roche | 8.38 | < 0.001 | 6.84 | < 0.001 | 6.69 | < 0.001 | 7.09 | < 0.001 |

CURB65: Confusion-Urea-Respiratory rate-Blood pressure-65-years-old index; HR: Hazard Ratio; p: p-value; N.S.: not significant; N.I.: not included; Low Ly.: low total lymphocyte count; IL-6: interleukin-6; LDH: lactate dehydrogenase; qSOFA: quick Sepsis related Organ Failure Assessment; TFS: Thermo Fisher Scientific. Model 1 adjusted by age, sex and Charlson index. Model 2 adjusted by age, sex, Charlson index and low total lymphocyte count. Model 3 adjusted by age, sex, Charlson index and high IL-6. Model 4 adjusted by age, sex, Charlson index and high LDH.

## DISCUSSION

Many efforts have been made since the first COVID-19 outbreak in China to identify poor prognosis factors in this worldwide and evolving pandemic. Several biomarkers such as low total lymphocyte count, high LDH serum levels, increased acute phase reactants (C-reactive protein, ferritin, fibrinogen, among others) or increased IL-6 serum levels have been proposed [6,15]. Recently, detection of SARS-CoV-2 RNA in blood has been suggested as a potential severity biomarker [12,13,16–18]. In this regard, our results reinforce these previous data with a main contribution to the management of COVID-19: we have determined a semiquantitative threshold for SARS-CoV-2 RNA detection in serum early after admission that allows establishing RNA values (“relevant CoVemia”) associated with higher mortality risk. Furthermore, we have shown that, technically, this finding is reproducible and, it is the most useful biomarker in the clinical setting for predicting mortality in COVID-19 patients.

From a technical point of view, we were able to detect CoVemia with high accuracy and concordance by using two commercially available kits, from Roche and TFS, as previously described [19]. These kits had been previously approved for detecting SARS-CoV-2 in nasopharyngeal samples. Although the sensitivity of TFS technology was slightly better for SARS-CoV-2 RNA detection in serum, this difference disappeared when the specific Ct cut-offs for each kit were applied to define relevant CoVemia. These results support the notion that the CoVemia detection can be highly reproducible since the Roche and TFS technologies are based on different procedures for RNA extraction and retrotranscription, as well as different genes for SARS-CoV-2 detection. Furthermore, our results validate previous reports that used the Roche kit and an “in-house” kit for SARS-CoV-2 RNA detection [12,13].

From a clinical point of view, we have shown that CoVemia has a high consistence as a severity biomarker in COVID-19, since it correlates with several variables that have been proposed to be associated with poor evolution in COVID-19, namely old age, comorbidity, qSOFA and CURB-65, as well as with laboratory markers such as high IL-6 or LDH serum levels and severe lymphopenia. Furthermore, multivariable Cox regression analysis identified high LDH serum levels and relevant CoVemia as independent and solid predictors for mortality after adjusting by age, sex, and presence of comorbidity. Neither qSOFA, CURB-65, low total lymphocyte count, nor high IL-6 serum levels maintained statistically significant association with mortality. Therefore, presence of relevant CoVemia was proved to be the best mortality predictor in our population since it provides a hazard ratio three times higher than that of the other significant variable, high LDH serum level.

Certain immunological mechanisms may underlie some of these associations. First, it is tempting to suggest that a decrease in activity or number of immune cells involved in lysis of infected cells may lead to higher viral replication and viral spread in the blood. In this regard, the correlation between IL-6 levels and CoVemia levels could be related with the recent description of lower cytotoxic potential of NK cells and senescence of CD8+ T lymphocytes associated to high IL-6 levels, which can be restored by tocilizumab treatment [20]. Second, association between lymphopenia and CoVemia could be related with lymphocyte viral infection and cellular lysis by cytotoxic cells. However, lymphopenia is more likely to be related with increased cellular migration into the lungs, as we have recently described for dendritic cells [21].

Strengths of our research include: immediacy of results during the emergency situation of the COVID-19 first wave, sufficient sample size (n = 193), fulfillment of STROBE standards for observational research, and a multifaceted view of patients including Immunology, Microbiology and many clinical assessments. However, this study has some limitations. Firstly, a single-center and retrospective design and, therefore, variability on the day of sample extraction. Nevertheless, the average time from beginning of symptoms to sample extraction (median of two days) was equal in patients with positive and negative CoVemia. Secondly, the moderate specificity of relevant CoVemia at admission to predict mortality. This finding could be associated to the variability over time of SARS-CoV-2 RNA detection in serum in COVID-19 patients [22]. Because of this variability, CoVemia detection at a specific time point may not properly reflect the state of viral infection, thereby influencing the assessment of severity outcomes. This could explain why previous descriptions of SARS-CoV-2 RNA detection in blood provided heterogeneous information on its value as severity biomarker [13,14,17]. Therefore, sequential assessment of CoVemia may be needed to improve its specificity to predict mortality. Last but not least, our findings are restricted to the so-called “first wave”, when an overloaded health system could not cover all the needs to provide critical care in some severity ill patients. Although the results are solid, studies in new waves experienced elsewhere are needed to validate their utility throughout the pandemic.

In summary, the study presented here has established the usefulness of SARS-CoV-2 RNA detection in blood in the initial assessment of patients admitted for COVID-19 due to its capability to predict mortality. This assessment would be easily implemented since it is reproducible, regardless of the commercially available kit used for SARS-CoV-2 RNA detection. Accordingly, information from tests widely used for diagnosis could be readily used for prognosis evaluation. However, the need for quantitative standardization of relevant CoVemia and evaluation of the meaning of persistent CoVemia must be addressed in further longitudinal studies.

## METHODS

### *Study design, population and data* collection

This is a retrospective observational study of patients admitted to Hospital Universitario La Princesa (HUP) during the early weeks of the first wave of COVID-19 outbreak in Spain (Flow chart in S3 Figure).

**S3 Figure:**
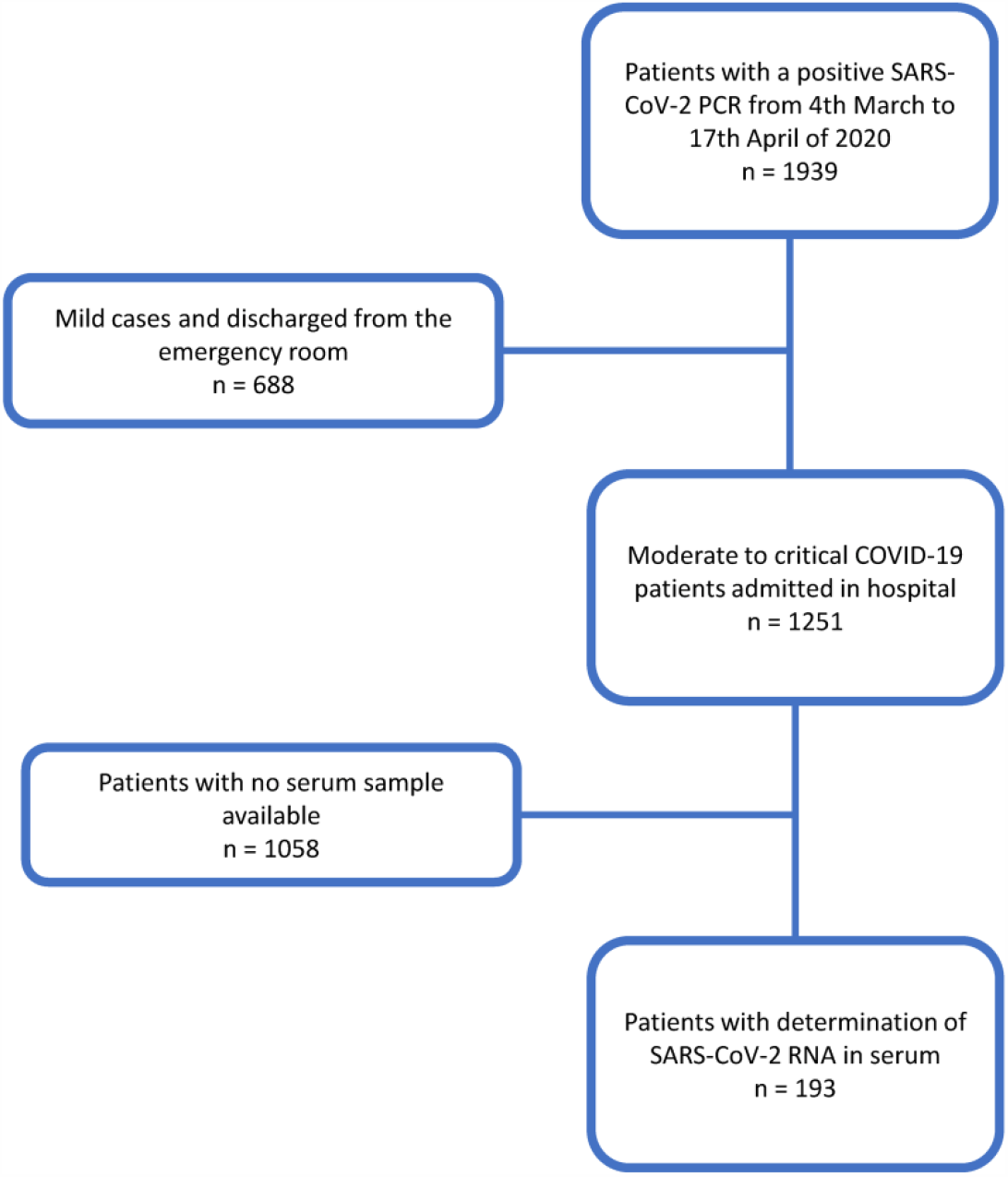
Flowchart of patients included in the study.

Study inclusion criteria were: a) patients with confirmed detection of SARS-CoV-2 RNA in nasopharyngeal and throat swabs (NPTS), and b) availability of serum samples at the Microbiology Department for additional measurement of SARS-CoV-2 RNA in blood. Regarding the last criterion, on average, serum samples were obtained at a median of two days from admission (interquartile range [IQR] 1 to 3) which nearly corresponded to one week after the beginning of symptoms (median: 8 days, IQR: 6 to 11).

Clinical, laboratory and therapeutic data were collected from electronic clinical records and included in an anonymized database as previously described [6]. Baseline clinical data refer to those obtained at admission day and baseline laboratory data refer to one to three days from admission date.

### SARS-CoV-2 RNA detection

Nasopharyngeal and throat swab samples were obtained and real-time reverse-transcription polymerase chain reaction (rRT-PCR) was performed with LightCycler^®^ Multiplex Virus Master (Roche Diagnostics, USA) using the LightMix^®^ Modular SARS and Wuhan CoV E-gene primers (Roche Diagnostics, USA), targeting the *e* gene as a first-line screening, followed by confirmatory testing with TaqPath™ COVID-19 CE-IVD Kit RT-PCR (Applied Biosystems™, USA) which detects three specific SARS-CoV-2 genomic regions: *orf-1ab, s*, and *n* genes. Both procedures were performed according to manufacturers’ directions on a Quant Studio-5 Real-Time PCR System (Applied Biosystems™, USA) and amplification curves were analyzed with QuantStudio™ Design and Analysis software (Applied Biosystems, USA), for their interpretation by a clinical microbiologist.

To evaluate CoVemia, we used 400 µL of surplus sera from routine HIV and hepatitis serology assessment that are usually frozen at −20 °C for internal quality controls. Serum samples were treated previously for virus inactivation and then tested with two rRT-PCR methods: cobas^®^ SARS-COV-2 test (Roche Diagnostics, USA), a qualitative assay for detection of SARS-CoV-2 RNA, and TaqPath™ COVID-19 CE-IVD RT-PCR Kit (Thermo Fisher Scientific [TFS], USA), a multiplex RT-PCR assay for qualitative detection of SARS-CoV-2 nucleic acids, as previously described [19]. The number of amplification cycles needed for the fluorescence signal to cross the threshold (Ct) was recorded. Briefly: a) Roche test detects a fragment of the *orf-1ab* region, specific of SARS-COV-2; and a conserved region of *e* gene, a structural envelope gene, for pan-sarbecovirus detection. Test was performed by cobas^®^ 6800 System (Roche Diagnostics, USA); an automatic platform for nucleic acid extraction and RT-PCR amplification and detection. Serum samples were processed according to manufacturer’s indications, following the same protocol used for SARS-CoV-2 detection in respiratory samples. Results were analyzed and interpreted automatically by the cobas^®^ 6800/8800 Software version 1.02.12.1002. B) TFS test detects three specific SARS-CoV-2 genomic regions: orf-1ab, s, and n genes. This technique requires a previous nucleic acid extraction from the sample, which was performed by the automatic eMAG^®^ Nucleic Acid Extraction System (Biomerieux, France), a validated system for the extraction of nucleic acids from serum and plasma samples. Extraction was carried out according to eMAG^®^ manufacturer’s directions, obtaining purified nucleic acids in 60 µL of elution buffer. Assay was performed using 5 µL of the eluted and according to the manufacturer’s instructions, by a QuantStudio™ 5 Real Time PCR System (Applied Biosystems, USA). Amplification curves were analyzed with QuantStudio™ Design and Analysis software version 2.4.3 (Applied Biosystems, USA). Interpretation of results was carried out by a clinical microbiologist, through amplification curve analysis.

### IL-6 serum level measurement

IL-6 serum levels were quantified in duplicate with the Human IL-6 Quantikine high sensitivity enzyme-immune assay from R&D Systems Europe Ltd. (Abingdon, UK), as described previously [6].

### Variables

We used two different variables to evaluate CoVemia: a) dichotomic variable: CoVemia was considered positive when at least one of the genes studied was positive according to the output of the two systems used; and b) quantitative variable: when more than one gene target was detected by one test, the mean Ct value of amplified genes was calculated. There was a high consistency among the results of the three genes analyzed with TFS and a slightly lower one for the two genes analyzed in the Roche system [19]. Since none of the two systems (Roche and TFS) provided positive results with Ct > 40, by consensus decision, we assigned Ct = 42 to those samples reported as negative (for all the genes) by the system.

Baseline IL-6 levels were considered high if > 30 pg/ml, according to their sensitivity and specificity to predict severity determined in a previous study [6]. In addition, in order to be able to compare different biomarkers, other continuous variables previously described as COVID-19 severity biomarkers were also transformed into dichotomic variables considering as cut-off point their median values in our population, namely: total lymphocyte count was considered low when < 700 cells/mm3, lactate dehydrogenase (LDH) serum level was considered high when > 400 U/L, qSOFA and CURB65 were considered high if > 1. Patients were considered elderly if age > 70 years.

Multiorgan dysfunction syndrome was considered when at least two organs reached a score > 1 in SOFA scale [23]. Multiorgan failure was considered when at least two organs reached a score > 3 in SOFA scale [24].

To analyze whether CoVemia could predict disease severity, two main outcomes were considered: need for ICU admission and all-cause in-hospital mortality. Only 14 patients out of 89 admitted to ICU did not require invasive mechanical ventilation.

### Statistical analysis

Statistical analyses were performed using Stata 14 for Windows (Stata Corp LP, College Station, TX, USA). Quantitative variables following a non-normal distribution were represented as median and IQR, and the Mann Whitney test was used to assess significant differences. Variables with a normal distribution were described by mean ± standard deviation (SD) and differences between groups were assessed with Student’s t-test. Qualitative variables were described as counts and proportions and Chi square or Fisher’s exact test was used for comparisons. Correlation between quantitative variables was analyzed using the Pearson correlation test. To estimate the 95% confidence interval (CI) of correlation coefficients we used the ci2 command of Stata. To evaluate the agreement between CoVemia assessed by Roche and TFS we estimated the intraclass correlation with the *icc* command of R.

To determine the variables associated with the need for ICU admission, we performed a multivariable logistic regression analysis that was first modeled by adding all the variables with a p-value lower than 0.15 to the bivariable analysis, namely low lymphocyte count, D-dimer, high LDH, low PaO2/FiO2 (< 250), COPD, high qSOFA, high CURB65, hypertension, C-reactive protein, and high IL-6 level. The final model was reached through backward stepwise removal of variables with p-value higher than 0.15 and using Wald tests to demonstrate that each model was better than its previous iteration. Once the final model was obtained, the dichotomic variables CoVemia (both Roche and TFS) were forced into the model in order to determine whether they were able to predict the need for ICU admission.

Odds ratio (OR) for mortality according to the presence of CoVemia was estimated for both systems with the *cs* command of Stata. Since differences between the two techniques were detected in the sensitivity to detect viremia and the capability of predicting mortality, receiver operating characteristic (ROC) analyses were performed using the *roctab* command in order to estimate the best cut-off point for each system. Cut-off values for relevant CoVemia were selected based on the best trade-off values between sensitivity, specificity and the percentage of patients correctly classified. Survival time was analyzed by Kaplan-Meier method with the *sts* command of Stata. Date of admission was considered the date of entry, and for exit date we considered the date of death. For those patients without this event, the last revision of the database (electronic chart or telephone call) on May 21st was used to censor their follow-up. Differences in time to death between different variables were analyzed by log-rank test.

To compare the predictive capability of baseline CoVemia with that of other baseline parameters (qSOFA, CURB65, total lymphocyte count, LDH and IL-6 serum levels) described in previous publications [12,13,25] we fitted different Cox regression models. In the first model after adjustment by age, sex and Charlson comorbidity index, the hazard ratio (HR) for all other variables including relevant viremia by TFS or Roche was estimated. Since qSOFA and CURB65 did not reach a p-value < 0.15, they were excluded from the analysis in the subsequent models. Model 2 was adjusted by the same variables included in model 1 and low lymphocyte count, the hazard ratio for the remaining variables was then estimated. Model 3 and 4 were developed as model 2 but substituting low lymphocyte count by high IL-6 and high LDH, respectively.

### Ethics

This study was approved by the local Research Ethics Committee (register number 4070) and it was carried out following the ethical principles established in the Declaration of Helsinki. All included patients (or their representatives) were informed about the study and gave an oral informed consent (registered in the electronic clinical chart) as proposed by AEMPS (Agencia Española de Medicamentos y Productos Sanitarios, *The Spanish Agency for Medicines and Medical Devices*) due to the COVID-19 emergency [26].

This article was written following the Strengthening the Reporting of Observational Studies in Epidemiology (STROBE) guidelines taking into consideration the difficulties to obtain all needed information in the setting of the COVID-19 pandemic (S1 Appendix)

## Supporting information

STROBE checklist

## Data Availability

All data will be fully available after acceptance.

## ABBREVIATIONS

CI: Confident interval
COPD: Chronic obstructive pulmonary disease
CoVemia: SARS-CoV-2 RNA in serum
COVID-19: Coronavirus disease-2019
CRP: C-reactive protein
Ct: Cycle threshold
FiO2: Fraction of inspired oxygen
HR: Hazard ratio
ICU: Intensive Care Unit
IL: Interleukin
IMV: Invasive mechanical ventilation
IQR: Interquartile range
IRR: Incidence risk ratio
LDH: Lactate dehydrogenase
NK: Natural killer
NPTS: Nasopharyngeal and throat swab
OR: Odds ratio
PaO2: Arterial oxygen tension
PCR: Polymerase chain reaction
qSOFA: Quick SOFA
RNA: Ribonucleic acid
ROC: Receiver operating characteristic
rRT-PCR: Real-time Reverse-Transcription Polymerase Chain Reaction
SARS-CoV-2: Severe Acute Respiratory Syndrome Coronavirus 2
SD: Standard deviation
SOFA: Sepsis related Organ Failure Assessment
TFS: Thermo Fisher Scientific

## ACKNOWLEDGEMENTS

Special thanks to our patients and relatives for agreeing with the use of pseudonymized clinical data and surpluses of clinical samples to perform this study, and PhD Manuel Gomez Gutierrez for his excellent editing assistance.

REINMUN-COVID group includes

Anesthesia: David Arribas Méndez, Rosa Méndez Hernández, Julia Hernando Santos, Mar Orts, Carlos Figueroa, Carlos Román, Antonio Planas.

Cardiology: Lourdes Domínguez Arganda, Pablo Martínez Vives, Guillermo Diego Nieto, Alberto Cecconi, Amparo Benedicto, Antonio Rojas González, Jesús Jiménez Borreguero.

Emergency Service: Carmen del Arco, Juan Mariano Aguilar, Natalia Villalba, Mónica Negro, Elvira Contreras, Ana del Rey, Cristina Santiago, Manuel Junquera, Raquel Caminero, Francisco Javier Val, Sonia González, Marta Caño, Isabel López, Andrés von Wernitz, Iñigo Guerra, Jorge Sorando, Lydia Chao, María José Cárdenas, Verónica Espiga, Alberto Pizarro.

Hematology Service: Rafael de la Cámara, Ángela Figuera Álvarez, Beatriz Aguado, Jimena Cannata, Javier Ortiz.

Hospital Pharmacy: Alberto Morell, Esther Ramírez, Amparo Ibáñez Zurriaga, María Pérez Abanades, Silvia Ruiz García, Tomás Gallego Aranda, María Ruiz, Concepción Martínez Nieto, José María Serra.

Immunology: Francisco Sánchez-Madrid, Cecilia Muñoz-Calleja, Arantzazu Alfranca, Ana Marcos-Jiménez, Santiago Sánchez-Alonso, Ildefonso Sánchez-Cerrillo, Laura Esparcia, Pedro Martínez-Fleta, Celia López-Sanz, Ligia Gabrie, Luciana del Campo Guerola, Elena Fernández, Reyes Tejedor.

Intensive Care Unit: Alfonso Canabal, Diego A. Rodriguez-Serrano, Nuria Arevalillo-Fernández, Marta Chicot Llano, Begoña González de Marcos, Pablo A. Patiño Haro, Marina Trigueros Genao, Begoña Quicios Dorado, David Jiménez Jiménez, Macarena Alonso González, Pablo Villamayor.

Internal Medicine-Infectious Diseases: Carmen Suárez, Ignacio de los Santos, José María Galván-Román, Emilia Roy Vallejo, Pablo Rodríguez-Cortes, Jesús Sanz, Eduardo Sánchez, Fernando Moldenhauer, Pedro Casado, Jose Curbelo, Angela Gutiérrez, Azucena Bautista, Nuria Ruiz Giménez, Angelica Fernández, Lucio García Fraile, Pedro Parra, Berta Moyano, Ana Barrios, Paloma Gil, Iluminada García Polo, Diego Real de Asúa, Beatriz Sánchez, Carmen Sáez, Marianela Ciudad, Marta Fernández Rico, Cristina Arévalo Román, Esperanza Morillo Rodríguez.

Medical Biology: Desiré Navas

Microbiology: Laura Cardeñoso Domingo, María del Carmen Cuevas Torresano, Diego Domingo García, Teresa Alarcón Cavero, Alicia García Blanco, Alexandra Martín Ramírez, María Auxiliadora Semiglia Chong, Ainhoa Gutiérrez Cobos, Nelly Daniela Zurita Cruz, Arturo Manuel Fraile Torres.

Pneumology: Julio Ancochea, Tamara Alonso, Pedro Landete, Joan Soriano, Carolina Cisneros, Elena García Castillo, Claudia Valenzuela, Francisco Javier García Pérez, Rosa María Girón, Javier Aspa, Celeste Marcos, Enrique Zamora, Ana Sánchez Azofra, Elena Ávalos Pérez-Urria, Gorane Iturricastillo, Mar Barrio Mayo, Encarna Rubia Garrido.

Rheumatology: Santos Castañeda, Rosario García-Vicuña, Isidoro González-Álvaro, Sebastián Rodríguez-García, Carlos Fernández-Díaz, Irene Llorente Cubas, Eva G. Tomero, Noelia García Castañeda, Ana Ma Ortiz, Cristina Valero, Miren Uriarte, Nuria Montes.

Surgery Department: Iñigo García Sanz, Francisco Eduardo Viamontes, Jesús Delgado Valdueza.

## Notes

### Competing Interest Statement

SCR-G reports grants from Spanish Rheumatology Foundation, during the conduct of
the study; nonfinancial support from Roche, Lilly, Pfizer, and Abbvie; personal fees and
nonfinancial support from Novartis, Sanofi, and MSD and from UCB-Pharma, outside
the submitted work.
JA reports grants and personal fees from GlaxoSmithKline and Boehringer Ingelheim;
grants from Linde Healthcare; and grants, personal fees, and nonfinancial support from
Roche and from Chiesi, outside the submitted work.
DAR-S reports personal fees from MSD, outside the submitted work.
RdC reports personal fees from MSD, ASTELLAS, Clinigen, Janssen, Roche, and
IQONE Health Care outside the submitted work.
RG-V reports grants, personal fees, and nonfinancial support from Abbvie, BMS, Lilly,
Novartis, Sanofi, Sandoz, and MSD; personal fees from Biogen and Celltrion and from
Mylan, outside the submitted work; personal fees and nonfinancial support from Pfizer;
grants from Roche; and grants and personal fees from Janssen.
CSF reports personal fees from Bayer, BMS, Daichi Sankyo, MSD, and Pfizer, outside the submitted work.
CM-C reports competitive grants from ISCIII during the conduct of the study.
IG-A reports grants from Instituto de Salud Carlos III, during the course of the study;
personal fees from Lilly and Sanofi; personal
fees and nonfinancial support from BMS and Abbvie; research support, personal fees,
and nonfinancial support from Roche Laboratories; and nonfinancial support from
MSD, Pfizer, and Novartis, not related to the submitted work.
The rest of the authors declare that they have no relevant conflicts of interests.

### Funding Statement

This study was funded with grants: Fondos Supera COVID19 by Banco Santander
and CRUE to CS, RG-V, CM and JA; RD16/0011/0012 and PI18/0371 to IGA, from
Spanish MINECO and Instituto de Salud Carlos III and co-funded by The European
Regional Development Fund (ERDF) A way to make Europe; and co-financed by the
Community of Madrid through the Covid 2019 Aid.
The work of ER-V has been funded by a Rio-Hortega grant CM19/00149 from the
Ministerio de Economia y Competitividad (Instituto de Salud Carlos III) and co-funded
by The European Regional Development Fund (ERDF) A way to make Europe. The
work of SCR-G has been funded by Fundacion Espanola de Reumatologia.
None of these sponsors have had any role in study design; in the collection, analysis,
and interpretation of data; in the writing of the report; and in the decision to submit the
article for publication.

### Author Declarations

This study was approved by the Research Ethics Committee of the Hospital Universitario de La Princesa, Madrid (register number 4070). All included patients (or their representatives) were informed about the study and gave an oral informed consent as proposed by AEMPS due to COVID-19 emergency.

